# RaBIt: An Effective Visualization-Driven Tool for Power and Sample Size Estimation in Two-Stage General Randomized Basket Trial Designs

**DOI:** 10.1101/2024.09.19.24313989

**Authors:** Desmond Zeya Chen, Sahil S. Patel, Aoqi Xie, Jiayin Chen, David Castle, Clement Ma

**Affiliations:** Centre for Addiction and Mental Health, Toronto, Ontario, Canada; Division of Biostatistics, Dalla Lana School of Public Health, University of Toronto, Toronto, Ontario, Canada; Genetics and Genome Biology Program, The Hospital for Sick Children, Toronto, ON, Canada; Department of Statistical Sciences, Faculty of Arts and Science, University of Toronto, Toronto, Ontario, Canada; Department of Statistics, North Carolina State University, Raleigh, North Carolina, United States; Department of Epidemiology, Biostatistics and Occupational Health, McGill University, Montreal, Quebec, Canada; Department of Psychiatry, University of Tasmania, Tasmania, Australia; Centre for Mental Health Service Innovation, Statewide Mental Health Service, Tasmania, Australia

**Keywords:** Shiny App, Basket Trial, Interim Analysis, visualization, novel trial designs, Adaptive designs

## Abstract

Basket trial designs with interim analysis have gained significant attention due to their adaptability, flexibility, and scalability. In response to the need for user-friendly tools that enhance the real-world applicability of these designs, we developed a web-based interface aimed at facilitating two-stage basket trial designs. Built using R Shiny, the tool was rigorously validated for output consistency by comparing it to an established R pipeline. Additionally, user testing was conducted to ensure the interface is intuitive and easy to use. The result is a freely accessible tool that provides effective and convenient visualizations for general basket trial designs with interim analysis, available at https://desmondzeyachen.shinyapps.io/AdaptiveTwoStageBasketTrialFeb14/. Future improvements may further expand the tool’s capabilities to accommodate the increasing complexity of trial designs needed by the research community.

## 1. Introduction

A master protocol trial is a state-of-the-art clinical trial design that innovatively aggregates the evaluation of multiple treatments or disease subtypes^1^. Its flexibility, adaptability, and effectiveness aided it gaining popularity since the onset of the millennium^2^. After the first master protocol managed the Imatinib Target Exploration Consortium Study B2225 basket trial design^3,4^, there has been a growing number of such trials being implemented^5,6^. Basket trials with biomarker enrichment have pioneered new frontiers in personalized medicine and thoroughly revolutionized the randomized trial design^7^.

Prior work from Chen et al^8^ has laid the statistical foundation for an adaptive basket trial design, it exquisitely addresses removal of given baskets based on interim results, while still accurately quantifies a trail-wide type 1 error rate, and a guidance for power. However, the impracticality of equal basket size assumption diminished its applicability. Such assumption is rarely fulfilled in real-word settings, considering the disproportionately distributed nature of disease subtypes prevalences among the population^9^. Following trial design guidance with such assumption may leads to a conservative, under-powered sample size estimate. After extensive investigation, we found an analytical solution to overcome this pitfall and developed a complementary software pipeline for efficient implementation and analysis^10^.

### 1.1 Study Objectives and Existing Work

We aim to provide trial researchers with support in proposal writing, trial monitoring, and trial execution. Majority of trial design software facilitating multiplicity through multi-arm multi-stage (MAMS) designs, only a few explicitly address basket trial designs. Berry Consultants developed a commercial software called FACTS^11^ (the “Fixed and Adaptive Clinical Trial Simulator”), which offers simulation-based analysis with highly flexible options across different trial classes. FACTS can simulate basket trial designs with early futility and interim analysis. However, sample size estimation was not supported, as researchers must search through the range of sample sizes manually. Among open-source packages and pipelines, the Integrated Platform for Designing Clinical Trials (https://trialdesign.org) currently features two basket design trial R pipelines based on Bayesian framework. One of these proposes a calibration to the existing Bayesian hierarchical approach^12^, significantly reducing type I error rate inflation compared to the uncalibrated version. The other, a Bayesian latent subgroup design^13^, clusters cancer types in basket trials into responsive and non-responsive subgroups, resulting in higher power and controlled type I error rates compared to standard approaches. Additionally, a frequentist pipeline that analytically solves for early futility of baskets is available in the supplemental section of Chen et al^8^. Although this overview highlights several essential tools and pipelines, it is important to recognize that there may be additional software options for basket trial design that are not included. Hence, to further enhance the software applicability and user experience, and to present a holistic elucidation of proposed trial design, we developed a web-based interactive R Shinny App^14,15^, to fill the methodological gap in two-stage general basket trial design^16^.

### 1.2 Target Audiences and Application Scope

Our tool is designed to support two-stage general randomized basket trials. Unlike conventional trials that focus on a single disease at a time, basket trials assess the efficacy of a single treatment across multiple homogenous diseases, significantly reducing the required sample size. The two-stage setup adds flexibility by allowing unresponsive disease baskets to be pruned at the interim stage, reallocating the remaining recruitment quota to responsive baskets. This early futility check ensures that resources are not wasted, improving the overall efficiency of the trial.

Initially developed for oncology studies, where it has been particularly effective in drug development for cancers and tumors, this design has become a widely adopted tool in various clinical research areas. Beyond oncology, the basket trial design is increasingly used in other therapeutic areas, including rare diseases and precision medicine.

The tool is particularly valuable for clinical trial researchers and biostatistical support teams due to its accessibility, intuitive usage, and convenience. Its user-friendly interface simplifies the complex process of designing and analyzing basket trials, making it easier for researchers to implement this advanced trial design without needing extensive statistical training.

## 2. Implementation

### 2.1 Statistical Methods

The null hypothesis for the design is that none of the baskets are active, while the alternative hypothesis is that only pre-specified baskets are active. As described by Chen et al^8^ and Patel et al^10^, the trial’s overall Type I error rate is calculated as the sum of rejection probabilities under the null hypothesis for different sets of baskets passing the interim analysis. The rejection probability is assessed based on whether all baskets pass through both the interim and final stages. Similarly, the power is estimated using the same sum of rejection probability function but under the alternative hypothesis, where the rejection region for pre-specified active baskets is adjusted according to the non-centrality parameter given by Chen et al^8^.

Analytically solving the sample size formula can be algebraically tedious, so we propose estimating it using a root-finding approach based on the power function. Unequal basket size ratios can be directly estimated from disease prevalence ratios, meaning that only the trial-wide sample size needs to be determined. Given the monotonic relationship between power and sample size, a recursive binary search^17^ algorithm is more efficient than computationally expensive iterative loops. This algorithm significantly improves the responsiveness of the app, making it faster and more efficient.

### 2.2 Variable Definitions and Input Parameters

Both calculations require the user to provide crucial trial characteristics (Figure 1) such as the standardized effect size (Δ) for each basket. Investigators can estimate this ratio based on subtype prevalence from published results. Additionally, the proportion of participants accrued at interim analysis (t) is needed. This is the cutoff point for interim analysis, expressed as a percentage of the total participants recruited, and should be consistent across all baskets. The interim significance level (αt) is also required, which is the designated α level for the interim analysis at proportion t. Any basket with a p-value exceeding this threshold will be dropped. The trial-wide significance level (α) must also be provided; this is the designated α level for the entire trial, representing the overall false positive rate.

**Figure 1:**
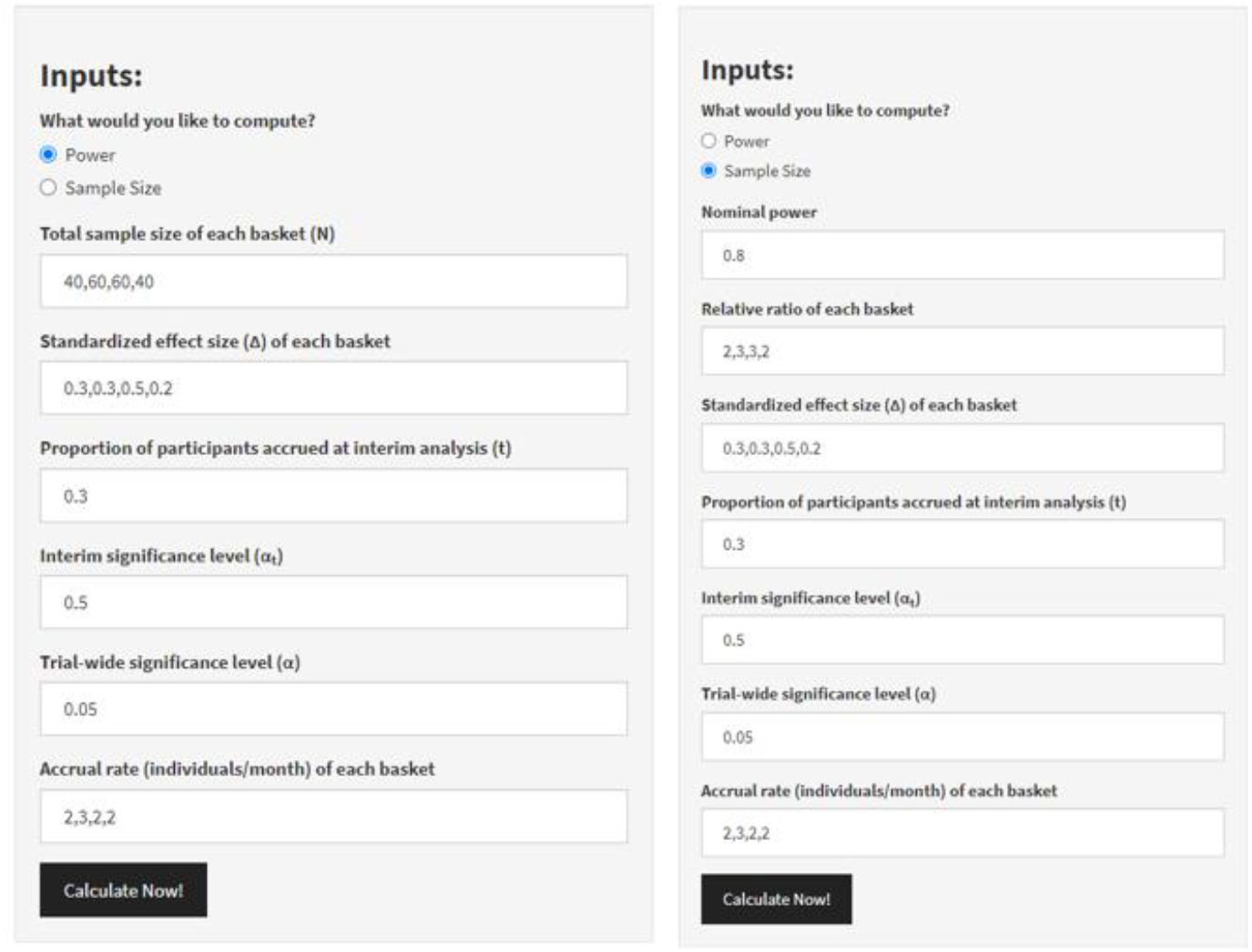
Input Parameters Required for Power or Sample Size Calculation

For sample size calculation, the sample size per basket is required. Conversely, for calculating the sample size per basket, the nominal power (designated 1−β level for the entire trial, or one minus the expected type II error rate) and the relative ratio of sample size in each basket must be provided. These are the primary parameters that directly influence the calculation, while the number of individuals recruited per month in each basket only affects the trial duration estimation.

### 2.3 System Architecture and User Interface Design

Having cloud computing power freely provided by the R Shiny^15^ server, the local machine is only responsible for capturing inputs and rendering outputs on the user interface, requiring minimal hardware resources. The remote server handles heavy-duty statistical computing tasks. The main tab of the tool consists of two sections: the input panel and the output panel. Power and sample size calculations are unified through an initial question that asks users which type of calculation they intend the tool to perform. The corresponding trial characteristics are then required for the calculation of their choice. On the output panel, an interactive trial design plot is displayed above the text summary. If incorrect inputs are provided, error messages will appear, masking all outputs. Additionally, a ‘Readme’ tab is available, offering an example, detailed explanations of the interface, and the statistical methods used, helping users gain hands-on experience.

### 2.4 Operational Workflow and Error Detection

Users can input trial characteristics with just a few clicks and text entries. The app is initiated by clicking the ‘Calculate Now!’ button. All input parameters are sent to the backend, where they are checked for missing data or syntax errors before being passed into the function. The function then verifies that the sample sizes are whole numbers within baskets at each stage. If any errors are found, they are promptly flagged for correction. Once all error checks are passed, the power or sample size is computed. Both summary statistics and trial characteristics inputs are compiled into a plot to visually represent the proposed trial. A text summary of the plot is displayed below for additional clarity. Users could then download the plot and text summary for their intended use.\

### 2.5 Validation, Reproducibility, and User Testing

To enhance reproducibility, we conducted extensive validation analysis and user testing. App’s outputs were systematically compared with results generated by the original R scripts used in the underlying statistical calculations from Patel et al^10^. This cross-verification ensured that the app replicates the established methods accurately. A variety of edge cases were tested, including extreme values of parameters and atypical trial configurations, to confirm the app handles all inputs robustly and responsively. This is crucial for identifying any scenarios where the app will fail and imposing corresponding error checkers as countermeasures. Additionally, we carefully examined the app’s error reporting mechanisms to ensure they provide clear and actionable feedback to users. This included verifying that the app correctly identifies and flags incorrect or missing inputs before proceeding with calculations. Finally, we conducted user acceptance testing with potential end-users like clinicians and biostatisticians to validate the app’s functionality in real-world scenarios. Participants were asked to replicate an example provided in the questionnaire, specifically for testing reproducibility. Feedback from these tests was incorporated to refine the app’s interface and usability.

## 3. Results

Our two-stage general basket design visualization tool is hosted at (https://desmondzeyachen.shinyapps.io/AdaptiveTwoStageBasketTrialFeb14/).

To ensure broad accessibility, the app is designed to be fully responsive, allowing seamless operation on desktops, tablets, or even smartphones. The intuitive user interface (Figure 2) is built to cater to users with varying levels of statistical knowledge, ensuring that even those unfamiliar with advanced statistical methods can easily navigate the tool. The interactive trial design figure can be easily downloaded as a JPEG file, and the design summary provides a clear, technical explanation of each characteristic and the summary statistics of the proposed trial design.

**Figure 2:**
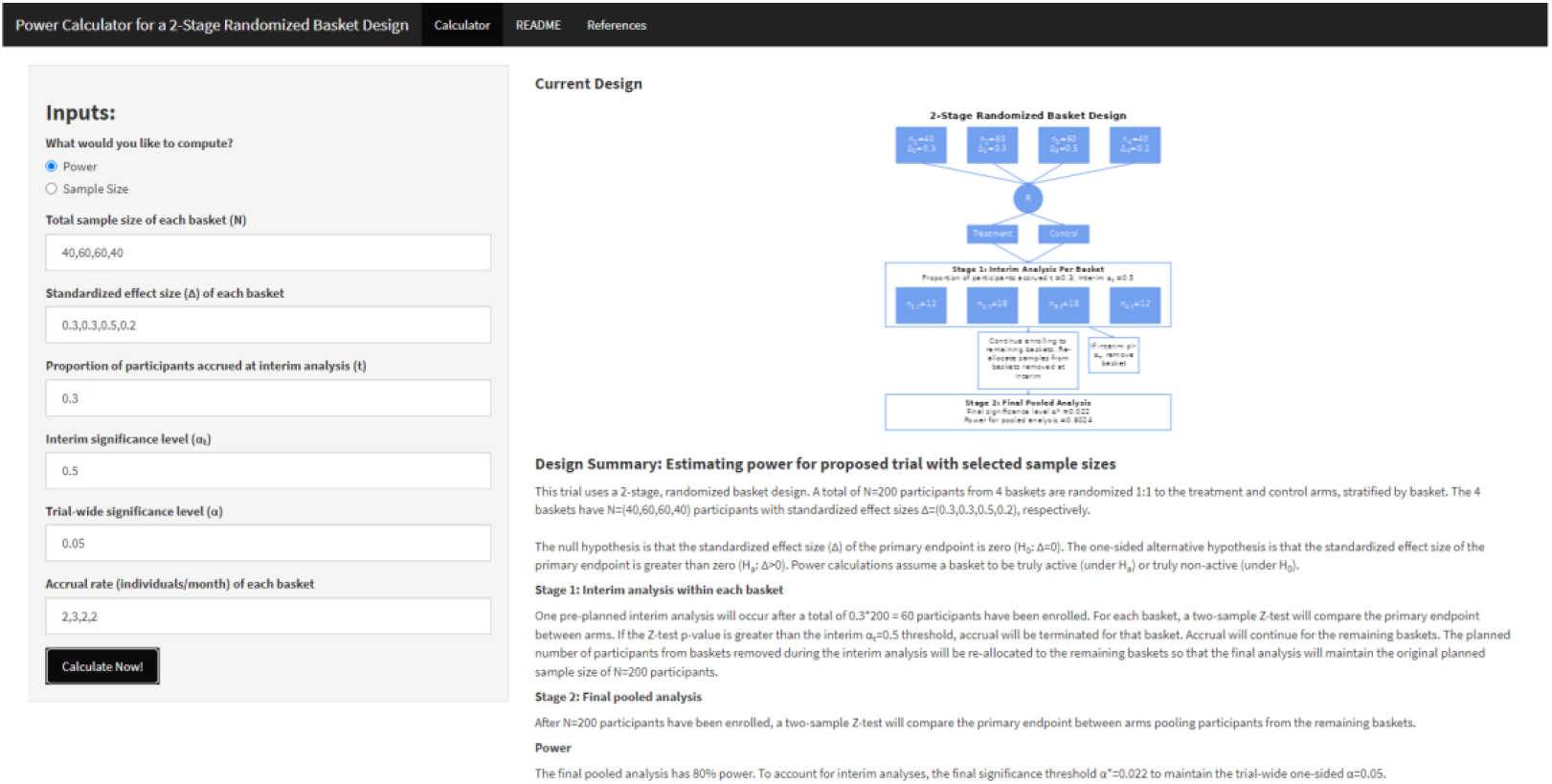
App Interface

### 3.1 User Interaction Feedback and Usability

Three users participated in the testing of the Shiny app, their feedback focused on ease of use, functionality, layout, and performance. All users found the app easy to navigate, especially with the help of default examples and detailed guidance provided in the Readme tab. Our original goal was to make the tool accessible to users with limited statistical training, aiming to simplify clinical trial design. However, one user noted that the use of highly specialized statistical language in the input variable names and explanations could create barriers for our target audience, potentially undermining our objective. We addressed this issue by incorporating additional explanations for each input from a trial design perspective, clarifying ‘What does this mean in clinical trial terminology?’ and ‘How could you decide on a value for it?’

The app was also presented at an internal meeting, where feedback related to sample size estimation functionality was raised. In response, we enhanced the app by adding features that support sample size estimation directly within the tool, further improving its utility for users designing clinical trials.

The app’s layout and functionality were deemed logical and well-organized, with users appreciating the clear instructions and intuitive design. Suggestions for enhanced result explanations and improved result visibility were carefully incorporated. In terms of performance, while the app generally worked well, users did experience occasional freezing or slower performance when working with larger sample sizes. These issues are common with the free version of the Shiny App server, and we plan to address them by either hosting the app on an internal server or upgrading to a premium account. All users expressed their willingness to use and recommend the app, citing its user-friendly interface and practical features. The user testing process, along with the feedback from the internal meeting, significantly improved the app’s usability and provided valuable, unbiased feedback from potential users.

### 3.2 Comparative Analysis

Primary trial characteristics like the total sample size, interim sample size, and number of baskets are influenced by the interrelated inputs: the number of baskets, sample size per basket, and interim proportions. Selecting an interim alpha of 0.3 allows for effective interim analysis without being overly conservative during partial recruitment. The trial-wide alpha is set at the conventional two-sided significance level of 0.025. We validate the Shiny App by comparing it against an R pipeline, using selected combinations of these inputs while controlling for effect sizes and alphas (Table 1).

**Table 1:**
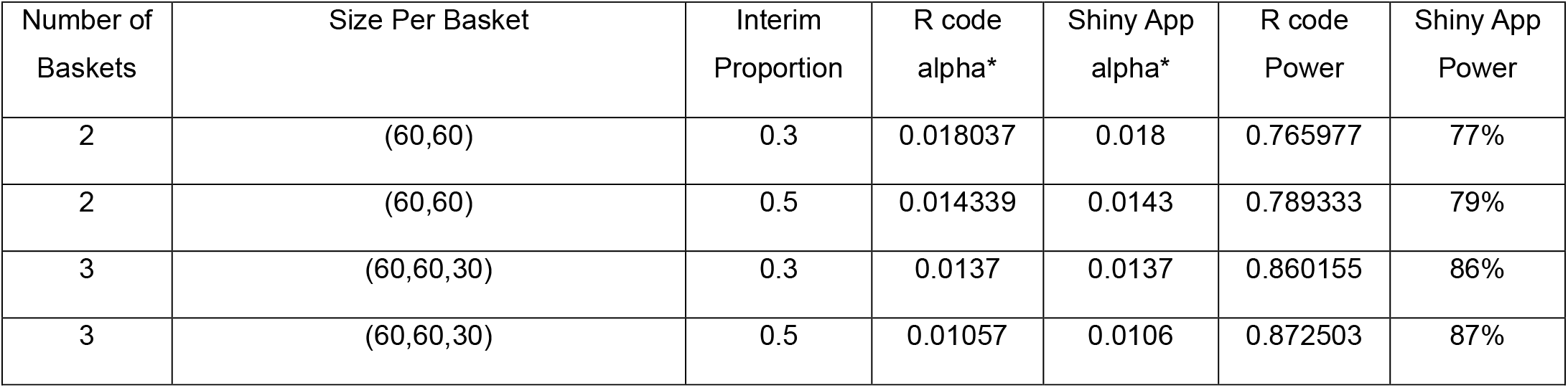
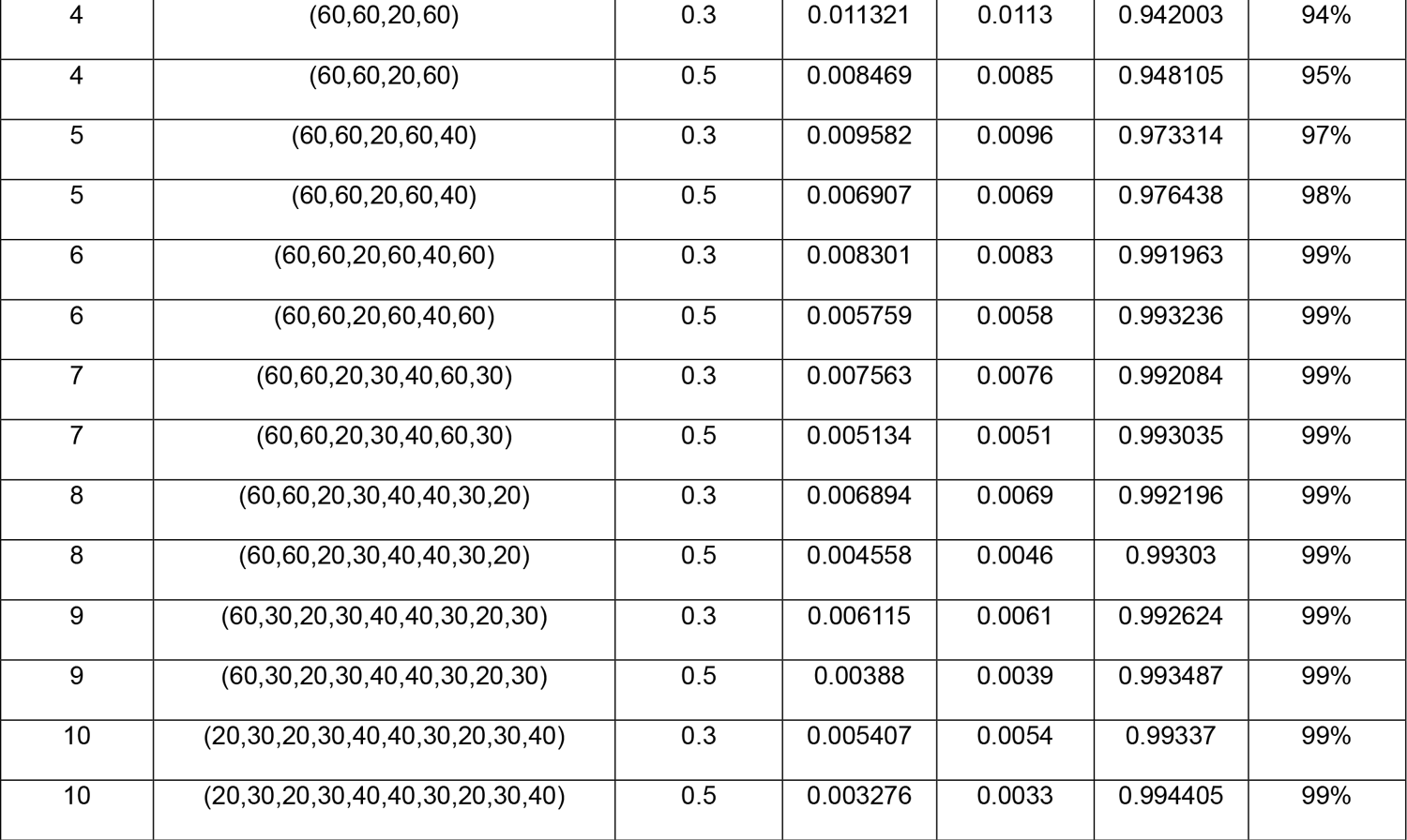
Comparative Analysis of the Tool vs R pipeline varying number of baskets and interim proportion. Assuming standardized effect size are all 0.5, all baskets are active interim alpha set at 0.3, and trial-wide alpha set at 0.025, alpha* is the recommended significance threshold of final analysis based on user provided trial wide alpha.

The alternative hypothesis is defined by user-specified effect sizes, with a value of 0 indicating that a basket will be pruned during interim analysis. To validate the tool under different alternative hypothesis scenarios, we varied the effect sizes per basket while keeping the sample size, number of baskets, interim proportion, and alphas constant (Table 2).

**Table 2:** Comparative Analysis of the Tool vs R pipeline varying effect size per basket. Varying effect size & whether the basket is truly active of (60,60,30). Interim proportion set at 0.3, interim alpha set at 0.3, and trial-wide alpha set at 0.025. Truly active vector, the corresponding baskets that pass interim analysis, deduced via effect size per basket that user provided.

| Table 2: Comparative Analysis of the Tool vs R pipeline varying effect size per basket |  |  |  |  |  |
| --- | --- | --- | --- | --- | --- |
| Effect Size Per Basket | Truly Active Vector | R code alpha* | Shiny App alpha* | R code Power | Shiny App Power |
| (0.2,0.2,0) | (1,1,0) | 0.01370037 | 0.0137 | 0.19221983 | 19% |
| (0.2,0.2,0.2) | (1,1,1) | 0.01369794 | 0.0137 | 0.23586268 | 24% |
| (0.5,0.2,0) | (1,1,0) | 0.01370189 | 0.0137 | 0.55850379 | 56% |
| (0.5,0.2,0.2) | (1,1,1) | 0.01369941 | 0.0137 | 0.58211621 | 58% |
| (0.5,0.5,0) | (1,1,0) | 0.01370145 | 0.0137 | 0.78413515 | 78% |
| (0.5,0.5,0.2) | (1,1,1) | 0.01370213 | 0.0137 | 0.79281031 | 79% |
| (0.5,0.5,0.5) | (1,1,1) | 0.0137017 | 0.0137 | 0.8601982 | 86% |
| Table 2: Varying effect size & whether the basket is truly active of (60,60,30). Interim proportion set at 0.3, interim alpha set at 0.3, and trial-wide alpha set at 0.025. Truly active vector, the corresponding baskets that pass interim analysis, deduced via effect size per basket that user provided. |  |  |  |  |  |

The results from the tool matched those generated by the R code after rounding, confirming that the tool correctly interprets the inputs and performs equivalently to the R code. This validation demonstrates the tool’s robustness and reliability in replicating established statistical processes. The consistency between the tool and the R code suggests that users can confidently employ the tool for interim analysis and hypothesis testing in clinical trials.

### 3.3 Worked Example

Recent findings have established a link between biological sex differences in the FGF13 gene on the X chromosome and the development of Autism^18,19^. We hypothesize that biological sex differences in the FGF13 gene on the X chromosome may influence treatment response in Autism. To mimic this scenario, we simulate a study cohort divided into four groups: FGF13+ Males, FGF13-Males, FGF13+ Females, and FGF13-Females. Reflecting the potential for varying prevalence of FGF13+ mutations across sexes, we assign the following hypothetical group proportions: 30%, 30%, 10%, and 30%, respectively. Assuming a total sample size of 200, a 30% accrual rate, and an interim alpha of 0.3 for an interim analysis, this simulated trial would have a power of 94%. To maintain an overall Type I error rate of 0.025, the final significance threshold would be recommended to be 0.011 (Table 1).

## 4. Discussion

Our open-source calculator is compiled to run on browser while computing were done remotely at RShiny^15^ server. Both power and sample size calculation functions were integrated into the same interface. Instead of manually constructing the pipeline, calibrating inputs, rephrasing outputs so its human readable, and plotting the design; our visualization tool automates all above tasks with just few simple steps. It provides users with an integrated, statistics enriched trial design figure and detailed textual exposition of the trial design alongside statistical results. These outputs can be conveniently assimilated as presented.

This user-friendly approach not only saves time but also reduces the potential for human error in trial design, ensuring a higher degree of accuracy and consistency. The tool’s ability to produce ready-to-assimilate outputs enhances its practical value, allowing for immediate integration into trial planning and reporting.

As clinical research moves towards more complex trial designs, basket trials have become increasingly popular due to their efficiency in exploring the effects of interventions across multiple disease types. These designs allow clinicians to investigate multiple patient populations simultaneously, improving resource allocation and potentially accelerating the discovery of effective treatments. Our tool fits seamlessly into this evolving landscape, providing an accessible means for researchers to engage with these complex designs without the need for extensive statistical training. By supporting the development and execution of basket trials, the tool also holds potential for educational purposes, enabling clinicians and researchers to independently explore and visualize these innovative trial structures. This flexibility, combined with the potential to expand into multi-stage and multi-arm trial designs, underscores the importance of developing tools that can keep pace with modern research needs. Moreover, due to analytical challenges in quantifying correlations between basket level treatment effects homogeneity, the existing framework may yield conservative estimates for both sample size and final significance thresholds. However, it also illuminates a potential area for future methodological work.

As basket trial designs gain popularity for biomarker-informed treatment outcome analysis, the sex variable, could either interact with, or directly affecting biomarkers intensity to influence efficacy^20,21^. Despite the prominence of biological sex-differentiated disease etiology in medical research^22^, limited attention has been given to incorporating the sex variable as the primary basis for basket grouping or post-trial outcome analysis^23^. We presented an emulating example of how basket trial could be utilized to disentangle the sex varied therapeutic effect.

Additionally, as the use of basket trials continues to expand, our tool is positioned to evolve alongside these developments, with potential future enhancements to support multi-stage designs, multi-arm trials, and other innovative trial methodologies. Gathering user feedback will be critical in guiding these developments to ensure the tool continues to meet the evolving needs of the research community.

## Data Availability

All data produced in the present work are contained in the manuscript

https://desmondzeyachen.shinyapps.io/AdaptiveTwoStageBasketTrialFeb14/

## Author Contributions

D.Z.C. and C.M. conceptualized the study. C.M. supervised the study. D.Z.C. and C.M. drafted the manuscript. D.Z.C. performed the analyses, developed the visualization software, and summarized the results. S.S.P. developed the base R code. S.S.P., A.X., J.C., and D.C. contributed to the investigation by conducting user testing of the application. All authors reviewed and edited the manuscript.

## Declaration of interests

The author(s) declare no conflicts of interest.

## Acknowledgements

We would like to thank Marcos Sanches and the rest of the biostatistics core at CAMH for their insightful questions during the internal meeting, which played a key role in shaping the development of this application. We would like to acknowledge the support of the University of Toronto Fellowship, provided by the University of Toronto for funding this research.

## Software and Web Resources

The visualization tool was developed using R^14^ version 4.3.2. The implementation involved several R packages, including shiny^15^ (version 1.8.0), shinythemes^24^ (version 1.2.0), ggplot2^25^ (version 3.4.4), ggtext^26^ (version 0.1.2), and ggforce^27^ (version 0.4.1).

## Notes

### Competing Interest Statement

The authors have declared no competing interest.

### Funding Statement

This study was funded by University of Toronto Fellowship

### Summary of Updates

A worked example was added at section 3.3.

